# Efficacy and safety of phentermine/topiramate in adults with overweight or obesity: A meta-analysis and systematic review

**DOI:** 10.1101/2020.10.26.20215475

**Authors:** Xiang-Guo Lei, Chen Lai, Ziyi Sun, Xi Yang

## Abstract

**Objective:** To examine the association between phentermine/topiramate therapy and weight loss and adverse events in adults with overweight problems or obesity by meta-analysis and systematic review.

**Methods:** Medical Subject Headings (MeSH) and free-text terms related to phentermine/topiramate were selected to search for eligible trials in PubMed, Cochrane Central Register of Controlled Trials (CENTRAL), and EMBASE up to April 18, 2020. The quality of randomized controlled trials was evaluated by Cochrane risk-of-bias tool. Meta-analysis was performed using random-effect models. Our systematic review protocol, registered on PROSPERO (registration number CRD42020188324).

**Results:** Phentermine/topiramate therapy resulted in a weight loss of 7.73 kg (95% confidence interval [CI]: 6.60, 8.85) compared to placebo. For phentermine/topiramate subjects in different weight loss subgroups, the weight loss of subjects with ≥5%, ≥10%, and ≥15% baseline weight loss were 3.18 (95% CI: 2.75, 3.67), 5.32 (95% CI: 4.53, 6.25), and 5.65 (95% CI: 3.55, 9.01), respectively. Phentermine/topiramate reduced waist circumference, blood pressure, blood sugar levels, and lipid levels. The adverse effects associated with the treatment mainly included Dysgeusia, Paresthesia, Dry mouth.

**Conclusions:** Phentermine/topiramate reduced body weight and was well tolerated. However, it increased the risk of nervous system-related adverse events to a certain extent, but the symptoms are not serious. Long-term clinical and pharmacological studies are needed to understand the long-term efficacy and safety of phentermine/topiramate.

**Study Importance Questions:**

- Phentermine/topiramate was approved by the FDA as an anti-obesity drug. However, the European Medicines Agency refused marketing authorization for phentermine/topiramate owing to safety concerns.
- Phentermine/topiramate reduced body weight and was well tolerated. However, it remarkably increased the risk of nervous system-related adverse events.
- In this study, the efficacy and incidence of adverse events of phentermine/topiramate were further evaluated through meta-analysis to provide reference for the clinical use of phentermine/topiramate.

## Introduction

Obesity is a chronic illness characterized by inordinate accumulation of body fat (1). it is a group of common metabolic disorders caused by multiple factors such as genetic factors, neuropsychiatric factors, endocrine factors, environmental factors, and reduced physical activity. In 2005, 23.2% (937 million) of the global adult population were diagnosed as being overweight and 9.8% (396 million) as obese; the respective numbers of adults with overweight problems and obesity are projected to be 1.35 billion and 573 million individuals in 2030, without adjusting for secular trends (2). Obesity is one of the primary causes of metabolic syndrome, which is characterized by a battery of metabolic disorders, including glucose intolerance, insulin resistance, central obesity, dyslipidemia, and hypertension (3). Metabolic disorders are associated with an increased risk of cardiovascular disease, rheumatism, and cancer (3). Thus, obesity is anticipated to be the leading cause of mortality and morbidity (5).

Phentermine/topiramate is unique in that most clinically applicable preclinical studies have been performed in humans on the two individual components (phentermine and topiramate), with both drugs having been approved as monotherapies for decades. Phentermine was approved by the US Food and Drug Administration (FDA) in 1959 (6). Topiramate underwent development as early as 1979 and was approved in the USA as a marketed pharmaceutical in 1996 (7). In 2012, Phentermine and topiramate (in one capsule) was approved by the FDA for use in adults with a body mass index (BMI) ≥30 kg/m^2^ or BMI ≥27 kg/m^2^ with at least one weight-related comorbidity (8). Phentermine/topiramate was approved by the FDA as an anti-obesity drug in 2012, and there has been an increasing interest in its use, application, and side effects (9). In the obese/overweight adult population, several research groups conducted randomized controlled trials (RCTs) to investigate the efficacy and safety of phentermine/topiramate in patients over a 2.5-year study period (9-14).

Phentermine/topiramate administration resulted in weight loss, which was found to be statistically significant from the RCT data. Phentermine/topiramate is available in the USA, but the European Medicines Agency refused marketing authorization for phentermine/topiramate in Europe owing to safety concerns (15). The most common adverse effects of phentermine/topiramate are dry mouth, constipation, and paresthesia (9, 11).

Given the contribution to weight loss, the efficacy of phentermine/topiramate may differ with dose and time. The effect of phentermine/topiramate on waist circumference, fasting glucose, fasting insulin, HbA1c, systolic blood pressure, diastolic blood pressure, total cholesterol, high-density lipoprotein cholesterol, low-density lipoprotein cholesterol, triglycerides, adiponectin, and adverse effects of phentermine/topiramate needs to be investigated. Therefore, in our study, we aimed to assess the efficacy and safety of phentermine/topiramate through a meta-analysis of RCTs, with focus on the effects and safety of phentermine/topiramate for adults with overweight problems or obesity.

## Methods

### Search strategy

Keywords and MeSH terms were selected and searched in PubMed, the Cochrane Central Register of Controlled Trials (CENTRAL), and EMBASE (eligible up to April 18, 2020). Articles not in English were excluded, and NCT number were identified by researching ClinicalTrials.gov. The search strategy is presented in Table S1 and our systematic review protocol, registered on PROSPERO (registration number CRD42020188324).

### Inclusion criteria

The inclusion criteria were as follows: studies must have included 1. a population of adults with overweight issues or obesity, with overweight defined as a BMI greater than or equal to 25 kg/m2 and obesity defined as a BMI greater than or equal to 30 kg/m^2^ (16); 2. intervention with phentermine/topiramate monotherapy; 3. comparison with a placebo; 4. outcomes including weight loss, weight loss percentage, waist circumference, fasting glucose, fasting insulin, HbA1c, systolic blood pressure, diastolic blood pressure, total cholesterol, high-density lipoprotein cholesterol, low-density lipoprotein cholesterol, triglycerides, adiponectin, and adverse effects of phentermine/topiramate; 5. RCT design; and 6. a follow-up duration of at least 4 weeks; 7. adverse events at least included in two studies.

### Data extraction and quality assessment

Full-texts and supplementary materials meeting the eligibility criteria were retrieved for data extraction. The following data were extracted from the qualified studies reviewed: first author; year of publication; study design; sample size; patient characteristics; intervention (dose of phentermine/topiramate); comparison (placebo); follow-up time; and results (loss of weight, weight loss percentage, etc.).

Disagreements were resolved by discussion between two investigators. Risk of bias was assessed using the Cochrane Collaboration Risk of Bias Tool (17). Xiang-Guo Lei and Chen Lai independently checked the data for accuracy, and the end outcomes for consistency.

### Statistical analysis

The primary outcomes of the study were change in weight (kg) and weight loss percentage between baseline and weight loss in subjects receiving phentermine/topiramate for 56 and 108 weeks (It is an exploratory analysis, not pre-specified in the PROSPERO registration.) and adverse events. For continuous outcomes, we used weighted mean differences (WMDs), e.g. weight. For dichotomous data, the outcomes were expressed as risk ratios (RRs), e.g. adverse effects. The quality of the evidence is upgraded when the overall effect estimate is dramatic: i.e. large (RR >2 or <0.5) or very large (RR >5 or <0.2) for dichotomous outcomes. Changes in waist circumference, fasting glucose, fasting insulin, HbA1c, systolic blood pressure, diastolic blood pressure, total cholesterol, high-density lipoprotein cholesterol, low-density lipoprotein cholesterol, triglycerides, and adiponectin were included in secondary outcomes. The meta-analysis used a random-effects model and heterogeneity, rather than sampling error, among studies was evaluated by the I^2^ statistic. *P* < 0.05 was regarded as significant. Completer analysis was performed when intention-to-treat (ITT) data were unavailable. For fasting glucose, a conversion factor of 18 mg/dL to 1 mmol/L was adopted. For fasting insulin, a conversion factor of 6.965 pmol/L to 1 μIU/mL was adopted. Standard deviation was required to perform the meta-analysis. However, most of the studies included in this meta-analysis did not provide standard deviations, but only standard error or a 95% confidence interval. Therefore, we employed a formula to calculate standard deviations as outlined in the Cochrane Handbook; for comparing a single pair, we combined two or more active therapy groups (18).

Owing to the small number of included studies, we were not able to evaluate publication bias using funnel graphics. STATA (version 12; StataCorp, College Station, Texas, USA) was used to analyze all statistical data.

## Results

### Study selection and characteristics

After primary search from the database, 476 references in English were sorted out. In total, six studies (published from 2011 to 2014) met the predefined inclusion criteria (Figure 1). The sample size varied from 45 to 2487. The follow-up duration ranged from 28 to 108 weeks. Cochrane risk-of-bias tool was assigned to each item included for quality evaluation of RCTs (Table 1). No other studies by manual searching yielded such as UpToDate and Google Scholar. The features of the selected trials are shown in Table S2.

**Table 1.** The Cochrane risk-of-bias tool summary for studies included in the meta-analysis

|  | Random sequence generation (selection bias) | Allocation concealment (selection bias) | Blinding of participants and personnel (performance bias) | Blinding of outcome assessment (detection bias) | Incomplete outcome data (attrition bias) | Selective reporting (reporting bias) | Other bias |
| --- | --- | --- | --- | --- | --- | --- | --- |
| Allison 2011 | + | + | + | + | + | + | - |
| Aronne 2013 | ? | + | + | + | + | + | - |
| Gadde2011 | + | + | + | + | + | + | - |
| Garvey 2012 | + | + | + | + | + | + | - |
| Garvey 2014 | ? | ? | ? | ? | + | + | - |
| Winslow 2012 | + | + | ? | ? | + | ? | - |

**Figure 1.**
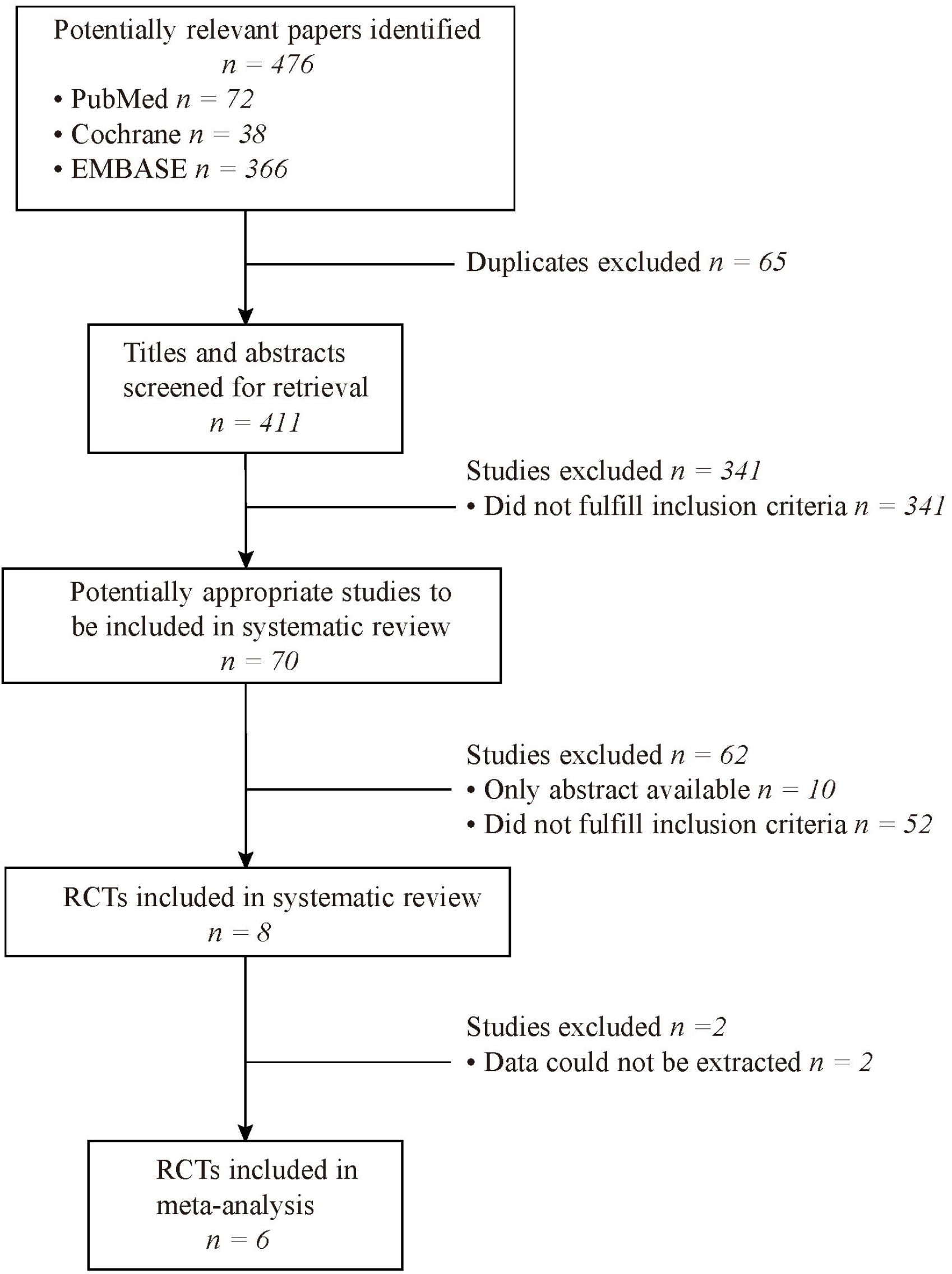
PRISMA flowchart for the identification and selection of studies for inclusion in the meta-review. RCT, randomized controlled trial.

### Methodological quality

Articles with significant shortcomings in study protocol or data analysis were excluded. All included studies met strict quality and eligibility criteria. All studies shown in Table S2, the combination of phentermine/topiramate and placebo was similar to the basic characteristics. Although the studies were double-blind, details of the randomization procedure were not mentioned in most studies.

### Primary outcomes

The subjects included in the study had an average estimated weight loss of 7.73kg (95% CI: 6.60, 8.85) after using phentermine/topiramate, and there was heterogeneity between studies (I^2^ = 82.1%) (Figure 2). Compared with the placebo group, the heterogeneity of the 3.75/23 mg phentermine/topiramate group could not be calculated because of the small number of studies included; the average estimated weight loss of the 7.5/46 mg group was 7.27 kg (95% CI: 6.40, 8.13), and no significant heterogeneity was detected (I^2^ = 35.8%). The average estimated weight loss of the 15/92 mg group was 8.35 kg (95% CI: 6.92, 9.79), and heterogeneity was detected (I^2^ = 87.5%) (Figure 3).

**Figure 2.**
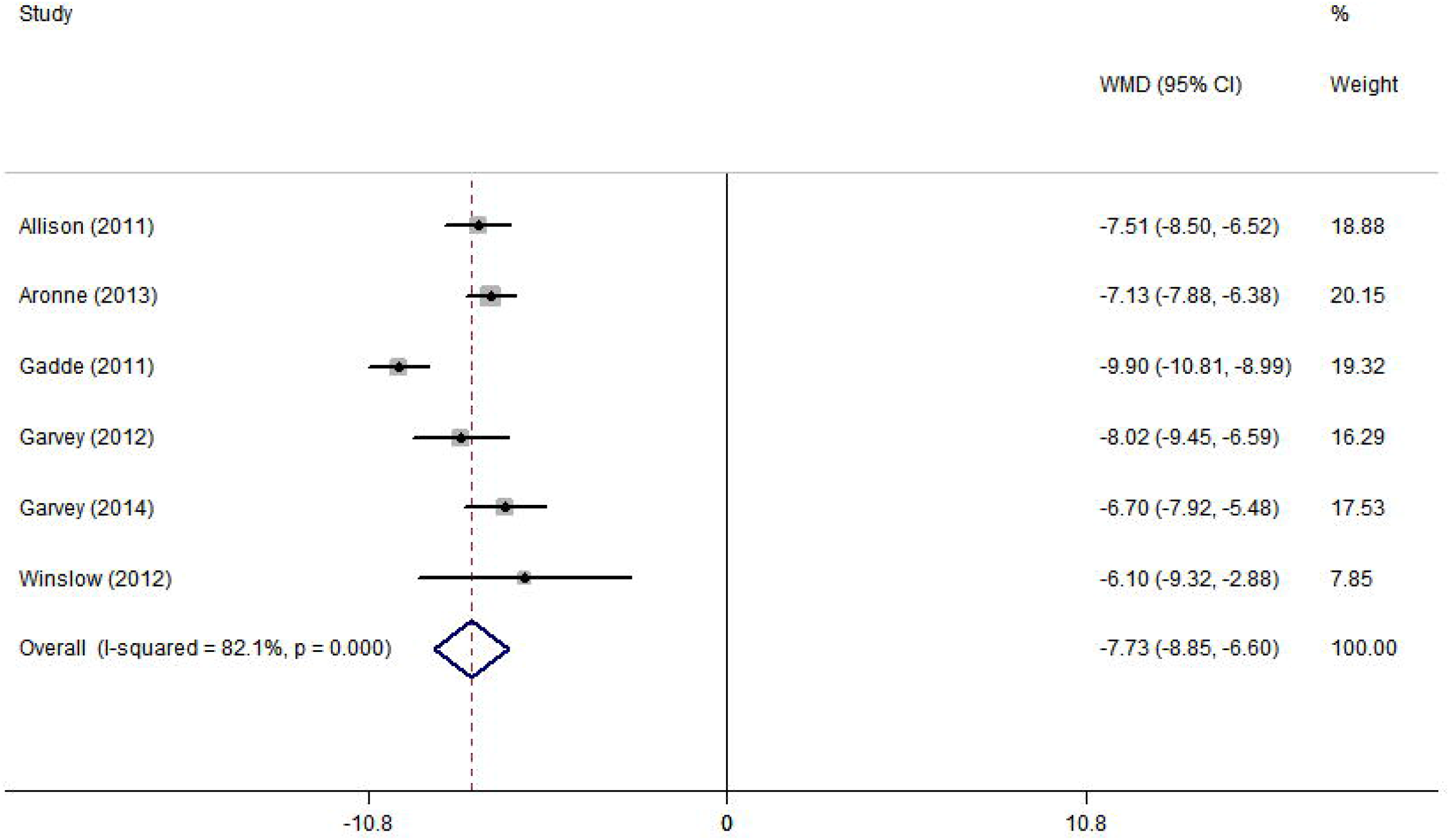
Average estimated weight loss of subjects included in the study. CI, confidence interval; WMD, weighted mean difference.

**Figure 3.**
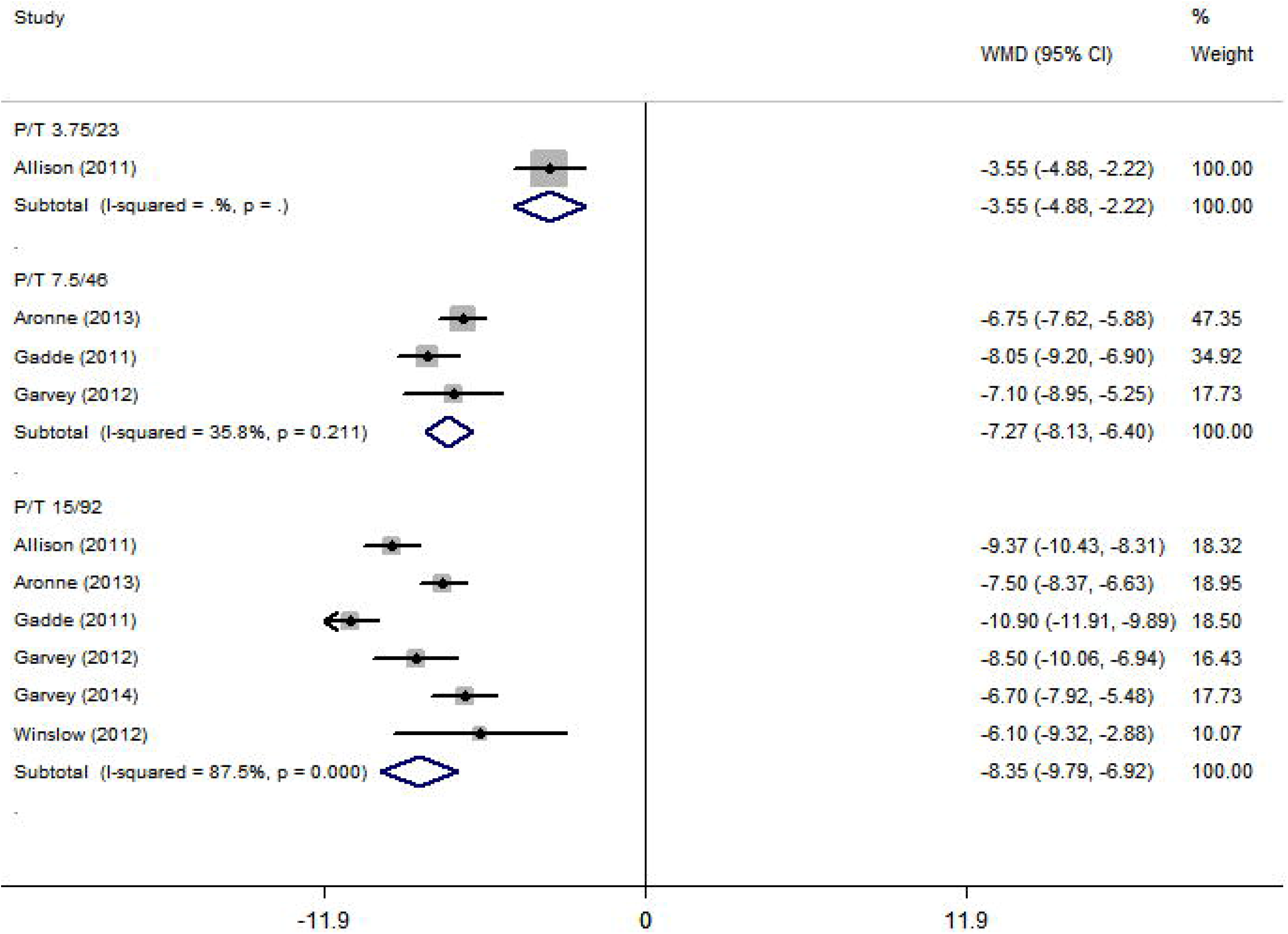
Average estimated weight loss of subjects receiving different doses of phentermine/topiramate. CI, confidence interval; WMD, weighted mean difference.

For subjects using phentermine/topiramate for 56 weeks, the average estimated weight loss was 8.07 kg (95% CI: 6.14, 9.99), and heterogeneity was detected (I^2^ =90.4%). For subjects who used the drug for 108 weeks, heterogeneity dropped significantly to 0 (Figure 4).

**Figure 4.**
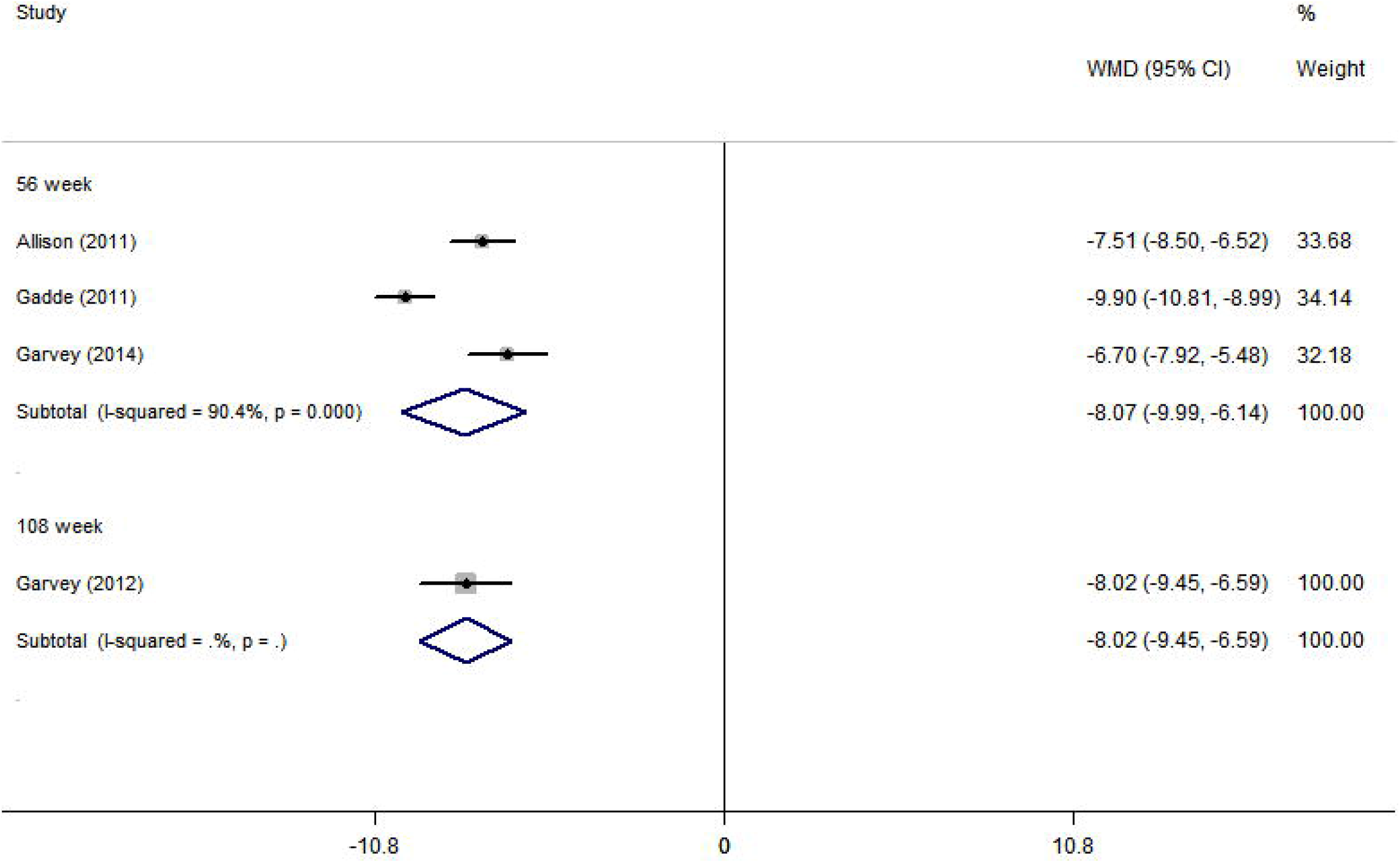
Weight loss in subjects receiving phentermine/topiramate for 56 and 108 weeks. CI, confidence interval; WMD, weighted mean difference.

For phentermine/topiramate subjects in different weight loss subgroups, compared with the placebo, the RR value of the number of subjects with ≥5% weight loss was 3.18 (95% CI: 2.75, 3.67), and heterogeneity was calculated (I^2^ = 47.1%). The RR value of the number of subjects with weight loss ≥10% was 5.32 (95% CI: 4.53, 6.25); however, heterogeneity significantly decreased to 0. The RR value of the number of subjects with ≥15% weight loss was 5.65 (95% CI: 3.55, 9.01), showing low heterogeneity (I^2^ = 43.2%). Weight loss ≥20% could not be combined analyzed as there were few studies (Figure 5).

**Figure 5.**
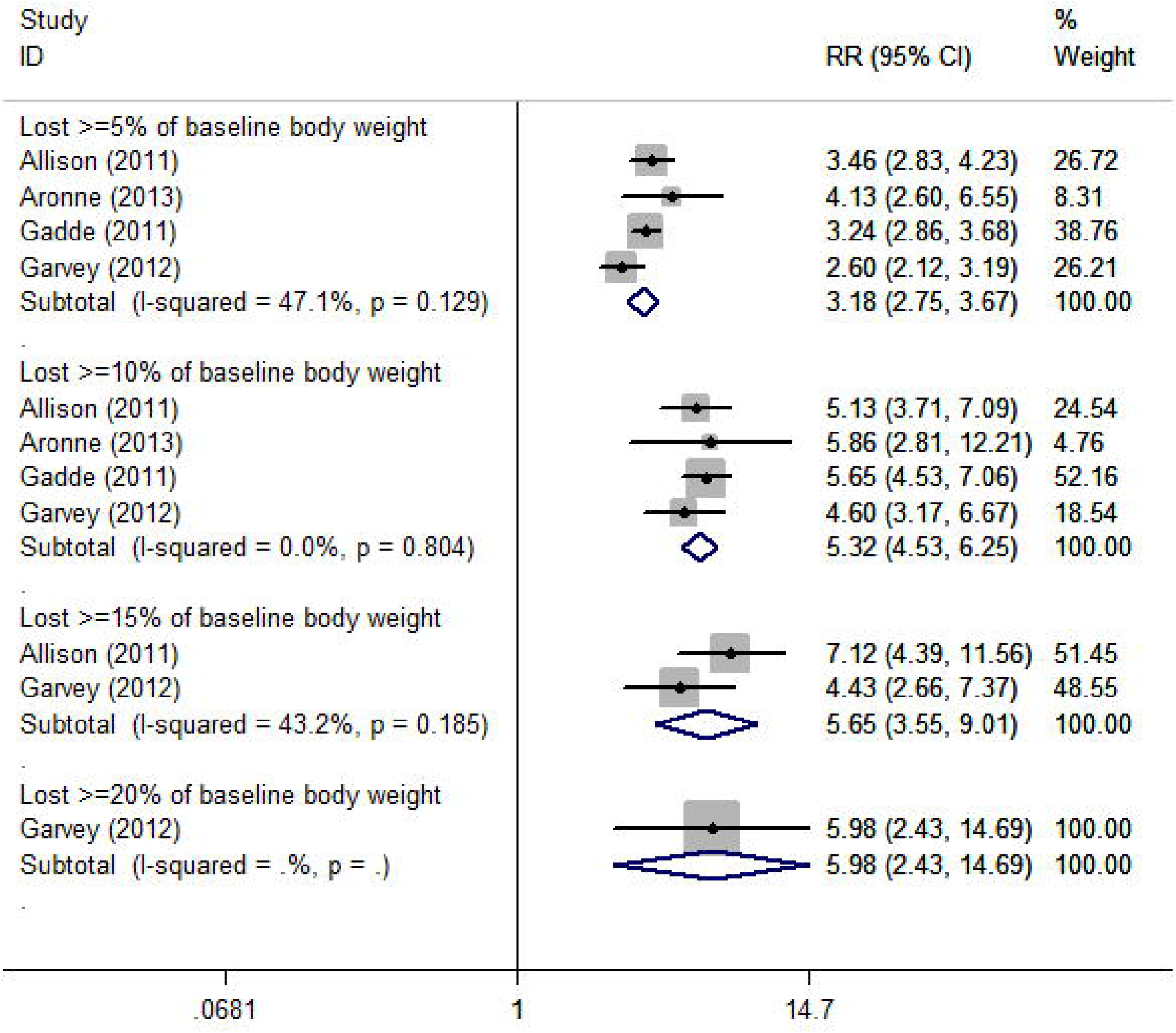
Comparison of the number of people taking phentermine/topiramate and placebo grouped by weight loss effect. CI, confidence interval; RR, risk ratio.

The adverse events associated with phentermine/topiramate treatment are presented in Table 2. The most common adverse events were Dysgeusia (RR = 8.86, 95% CI: 5.65, 13.89), Paresthesia (RR = 8.51, 95% CI: 6.20, 11.67), Dry mouth (RR = 6.71, 95% CI:5.03, 8.94), followed by Disturbance in attention (RR = 4.48, 95% CI: 2.39, 8.41), Irritability (RR = 4.10, 95% CI: 2.29, 7.33), Hypoesthesia (RR = 3.81, 95% CI: 1.32, 11.00), Constipation (RR = 2.43, 95% CI: 2.02, 2.93), Dizziness (RR = 2.26, 95% CI: 1.72, 2.98).

**Table 2.** The adverse events associated with phentermine/topiramate treatment

| Adverse event | Number of studies | Sample size (total, phentermine/topiramate, placebo) | RR | 95% Confidence interval |  |
| --- | --- | --- | --- | --- | --- |
| Dysgeusia | 5 | 5065, 3200, 1865 | 8.86 | 5.65 | 13.89 |
| Paresthesia | 6 | 5195, 3275, 1920 | 8.51 | 6.20 | 11.67 |
| Dry mouth | 6 | 5195, 3275, 1920 | 6.71 | 5.03 | 8.94 |
| Disturbance in attention | 3 | 4345, 2730, 1615 | 4.48 | 2.39 | 8.41 |
| Irritability | 3 | 4345, 2730, 1615 | 4.10 | 2.29 | 7.33 |
| Hypoesthesia | 2 | 1582, 1046, 536 | 3.81 | 1.32 | 11.00 |
| Constipation | 6 | 5195, 3275, 1920 | 2.43 | 2.02 | 2.93 |
| Dizziness | 5 | 5150, 3253, 1897 | 2.26 | 1.72 | 2.98 |
| Anxiety | 3 | 4152, 2591, 1561 | 1.88 | 1.24 | 2.86 |
| Depression | 3 | 4152, 2591, 1561 | 1.73 | 1.18 | 2.55 |
| Insomnia | 5 | 5150, 3253, 1897 | 1.67 | 1.34 | 2.08 |
| Vision blurred | 2 | 1860, 1238, 622 | 1.54 | 0.93 | 2.53 |
| Influenza | 4 | 2665, 1761, 904 | 1.42 | 1.03 | 1.96 |
| Nausea | 5 | 5150, 3253, 1897 | 1.41 | 1.11 | 1.79 |
| Urinary tract<br>infection | 3 | 3290, 2015, 1275 | 1.40 | 1.04 | 1.87 |
| Bronchitis | 5 | 5150, 3253, 1897 | 1.35 | 1.05 | 1.74 |
| Diarrhea | 6 | 5195, 3275, 1920 | 1.30 | 1.02 | 1.65 |
| Sinusitis | 5 | 5065, 3200, 1865 | 1.30 | 1.07 | 1.58 |
| Fatigue | 4 | 5020, 3178, 1842 | 1.24 | 0.96 | 1.59 |
| Back pain | 4 | 5020, 3178, 1842 | 1.23 | 0.98 | 1.55 |
| Cough | 2 | 1860, 1238, 622 | 1.13 | 0.70 | 1.83 |
| Nasopharyngitis | 6 | 5195, 3275, 1920 | 1.13 | 0.96 | 1.33 |
| Headache | 5 | 5150, 3253, 1897 | 1.07 | 0.90 | 1.27 |
| Upper respiratory<br>tract infection | 5 | 5065, 3200, 1865 | 1.06 | 0.93 | 1.22 |
| Arthralgia | 3 | 3290, 2015, 1275 | 0.84 | 0.65 | 1.10 |
| Gastroenteritis | 2 | 720, 470, 250 | 0.83 | 0.47 | 1.47 |

**Table 3.** Secondary outcomes of phentermine/topiramate treatment in individuals with obesity

| Secondary outcomes | Number of studies | Sample size (total, phentermine/topiramate, placebo) | WMD | 95% Confidence interval |  |
| --- | --- | --- | --- | --- | --- |
| Waist circumference (cm) | 3 | 3986, 2406, 1580 | -6.16 | -6.77 | -5.55 |
| Fasting glucose (mmol/L) | 6 | 4697, 2882, 1815 | -0.17 | -0.23 | -0.11 |
| Fasting insulin (μIU/mL) | 4 | 3174, 1945, 1229 | -4.21 | -5.77 | -2.65 |
| HbA1c (%) | 3 | 1078, 709, 369 | -0.14 | -0.22 | -0.06 |
| Systolic BP (mmHg) | 5 | 4527, 2745, 1782 | -2.92 | -3.75 | -2.09 |
| Diastolic BP (mmHg) | 5 | 4527, 2745, 1782 | -0.96 | -1.52 | -0.40 |
| Total cholesterol (%) | 3 | 3705, 2230, 1475 | -2.30 | -3.33 | -1.27 |
| HDL cholesterol<br>(%) | 4 | 4380, 2678, 1702 | 4.62 | 3.43 | 5.81 |
| LDL cholesterol<br>(%) | 3 | 4241, 2600, 1641 | -0.96 | -2.47 | 0.55 |
| Triglycerides (%) | 4 | 4380, 2678, 1702 | -13.38 | -16.37 | -10.39 |
| Adiponectin<br>(µg/mL) | 2 | 2267, 1443, 824 | 1.48 | 1.22 | 1.74 |

BP, blood pressure; HDL, high-density lipoprotein; LDL, low-density lipoprotein; WMD, weighted mean difference

### Secondary outcomes

The secondary outcomes of phentermine/topiramate and placebo treatments are presented in Table 2. Compared with the placebo group, the waist circumference of the phentermine/topiramate group decreased by an average of 6.16 cm (95% CI: 5.55, 6.77). The total average estimates of systolic and diastolic blood pressure reduction were 2.92 mmHg (95% CI: 2.09, 3.75) and 0.96 mmHg (95% CI: 0.40, 1.52), respectively. Fasting blood glucose and fasting insulin levels dropped by an average of 0.17 mmol/L (95% CI: 0.17, 0.23) and 4.21 μIU/mL (95% CI: 2.65, 5.77), respectively. The total cholesterol and triglyceride percentages in the phentolamine/topiramate group were significantly reduced, with average reductions of 2.30% (95% CI: 1.27, 3.33) and 13.38% (95% CI: 10.39, 16.37), respectively. A marked increase in high-density lipoprotein cholesterol was observed.

## Discussion

Meta-analysis of RCTs evaluating phentermine/topiramate use in adults with overweight problems and obesity showed that phentermine/topiramate use resulted in statistically significant weight loss compared with placebo. Within a certain range, the degree of weight loss was positively correlated with the drug dose; phentermine/topiramate was less effective for weight loss in 108 weeks of use, although most individuals were able to maintain the weight they lost in 56 weeks. Phentermine/topiramate treatment resulted in an average weight loss of 7.73 kg but also improved indicators that are beneficial to the cardiovascular system, such as waist circumference, blood pressure, blood sugar, and blood lipid levels. The most common adverse events associated with phentermine/topiramate were paresthesia, dry mouth, and constipation, followed by dysgeusia and dizziness; thus, phentermine/topiramate treatment seems to affect the nervous system. The placebo group performed higher in terms of withdrawal rates. This could be due to the failure to lose weight. Poor efficacy promoted the formation of adverse emotional responses in the placebo group, which possibly led to an even poorer efficacy. Phentermine is a sympathomimetic amine anorexia agent. There are insufficient animal and human studies combining phentermine with topiramate, and the mechanism of weight loss remains unclear. The current popular mechanisms of action of phentermine/topiramate mechanism are as follows: first, phentermine/topiramate may promote the release of neurotransmitters such as dopamine and norepinephrine, while feedback inhibits catecholamine release from the hypothalamus and reduces appetite by blocking catecholamine reuptake by the nerve endings. Topiramate inhibited the ionic receptor subtypes activated by kainate and glutamic acid, and some types of voltage-gated Na^+^ channels and Ca^2+^ channels. In addition, it regulates some types of K^+^ channels and GABA-A receptors, either positively or negatively depending on the receptor subtype (19). Second, phentermine/topiramate not only increases heat production and reduces energy efficiency, but also increases glucose uptake and the utilization of muscle, fat, and other tissues (9). Owing to the effect of phentermine/topiramate sustained-release agents on blood lipids and blood sugar, phentermine/topiramate may reduce the use of antihypertensive and lipid-lowering drugs.

### Comparisons with existing literature

Our data are in line with previous reviews (9-14) that used data published in clinical trial reviews to assess the efficacy and safety of phentermine/topiramate. These reviews concluded that phentermine/topiramate distinctly reduced body weight, but also distinctly increased the risk of adverse events.

### Comparison with orlistat and naltrexone/bupropion

Orlistat and naltrexone/bupropion are also FDA-approved weight loss drugs (20, 21). Phentermine/topiramate showed a stronger weight loss effect than orlistat and naltrexone/bupropion. Treatment with orlistat is associated with body weight reduction (WMD: −2.12, 95% CI: −2.51, −1.74, p < 0.001). Participants who were treated with naltrexone/bupropion lost significantly more body weight when compared with the baseline value (MD = −2.53 kg (−3.21 to −1.85). However, phentermine/topiramate resulted in a greater change in its WMD, which was 8.00 kg (95% CI: 7.17, 8.83). The most significant weight loss due to orlistat treatment occurred at 52 weeks. Nonetheless, weight loss from phentermine/topiramate gradually stabilized after 56 weeks (20, 21). Proper exercise and a healthy diet are required to maintain the weight loss effect; failure to incorporate these components can result in an obvious weight rebound. A meta-analysis on the efficacy and adverse reactions associated with diet pills showed that orlistat and naltrexone/bupropion were safer than phentermine/topiramate. In subjects with ≥5% weight loss, the odds ratios of individuals with adverse events associated with orlistat and naltrexone/bupropion were 2.69 (2.36 to 3.07) and 3.90 (2.91 to 5.22), respectively, compared with the placebo group, in contrast to an odds ratio of 9.10 (7.68 to 10.78) in the phentermine/topiramate group. For subjects with ≥10% weight loss, the odds ratios of individuals with adverse events associated with orlistat and naltrexone/bupropion were 2.41 (2.08 to 2.78) and 4.11 (2.80 to 6.05), respectively, compared with 11.34 (9.10 to 14.13) in the phentermine/topiramate group (22). When the weight loss proportion gradually increased, only orlistat showed a stabilization or decline in the risk of adverse events based on research data (19, 21, 22) evaluating several diet pills.

### Strengths and limitations

As phentermine/topiramate has been authorized by the FDA as a new drug recently, the number of RCTs using these drugs is not very large. Therefore, our meta-analysis provides researchers with directions to further evaluate the indications and contraindications of phentermine/topiramate. In addition, we provide a summary on the possible pharmacological mechanisms of this drug. However, there are certain limitations to our meta-analysis. First, there were few original studies on phentermine/topiramate, and only six RCT articles were eligible for analysis. Second, previously published studies have reported that the absolute weight loss effect of phentermine/topiramate in adults with overweight problems and obesity was statistically significant. However, it was difficult to detect the presence of a publication bias owing to a lack of literature, and we were unable to prepare funnel charts to obtain publication bias data. Third, all related studies were conducted in the USA and included only adults with overweight problems and obesity, limiting the generality of the results of the study. Finally, useable data showed that phentermine/topiramate had good safety and tolerability; only a few neurological symptoms require further research. However, we neglected to count the related adverse reactions of special population, such as drug-related miscarriages and malformed fetuses in maternity.

## Conclusions

Phentermine/topiramate treatment resulted in a moderate reduction of absolute weight in adults with overweight problems and obesity. It is likely to have a protective effect on the cardiovascular system because it effectively improved triglycerides, total cholesterol, blood sugar, blood pressure, and other indicators. Except for constipation and some minor neurological symptoms, phentermine/topiramate was well tolerated. Further research is needed on the dosage of phentermine/topiramate in different patient groups to understand its effect on weight management in individuals with obesity. Long-term clinical and pharmacovigilance studies are needed to understand the long-term efficacy and safety of phentermine/topiramate.

## Supporting information

manuscript files

manuscript files

manuscript files

manuscript files

## Data Availability

date available

## Acknowledgements

I would like to thank my friends, family, and teachers for their constant encouragement.

