## Supplementary material for "Efficacy and safety of phentermine/topiramate in adults with overweight or obesity: A meta-analysis and systematic review": manuscript files

### The effects and safety of phentermine/topiramate for overweight or obese adults: a meta-analysis

To enable PROSPERO to focus on COVID-19 registrations during the 2020 pandemic, this registration record was automatically published exactly as submitted. The PROSPERO team has not checked eligibility.

#### Citation

Xiang-Guo Lei, Chen Lai. The effects and safety of phentermine/topiramate for overweight or obese adults: a meta-analysis. PROSPERO 2020 CRD42020188324 Available from: https://www.crd.york.ac.uk/prospero/display_record.php?ID=CRD42020188324

#### Review question

To assess the effects and safety of phentermine/topiramate (PHEN/TPM) for overweight or obese adults.

#### Searches

A four-step search strategy is planned. Firstly, we will identify keywords and MeSH terms in PubMed. Secondly, the terms will be searched in PubMed, the Cochrane Central Register of Controlled Trials (CENTRAL) and EMBASE. Thirdly, randomized clinical trials analyzing use phentermine/topiramate in overweight or obese adults will be selected. Fourthly, references of included studies will be searched for additional papers.
(phentermine AND topiramate) OR (PHEN AND TRP) OR (PHEN AND TPM) OR VI-0521 OR Qsymia OR Qnexa

#### Types of study to be included

We will include randomized controlled trials

#### Condition or domain being studied

In the last 8 years, phentermine/topiramate (PHEN/TPM) have been introduced for obese therapy. It have been associated with decreased weight level. Abstract In 2012, the US Food and Drug Administration approved phentermine/topiramate for long-term weight loss. However, The European Medicines Agency refused marketing authorisation for phentermine/topiramate due to safety concerns. we focus on the effects and safety of phentermine/topiramate for overweight or obese adults.

#### Participants/population

overweight or obese adults

We will define overweight as a body mass index (BMI) : 25 kg/m-squared and < 30 kg/m-squared (AAFP 2013; WHO 2004).
Obesity will be defined by a BMI : 30 kg/m-squared (AAFP 2013; WHO 2004).

#### Intervention(s), exposure(s)

phentermine/topiramate

#### Comparator(s)/control

Placebo compared with phentermine/topiramate

#### Main outcome(s)

Weight loss (kg); The percentage of people who lost weight and Adverse events

#### * Measures of effect

weighted mean difference; relative risks

#### Additional outcome(s)

Waist circumference (cm), Fasting glucose (mmol/ L), Hb A1c (%), Fasting insulin (?IU/ mL), Systolic BP (mm/ Hg), Diastolic BP (mm/ Hg), Total cholesterol (%), HDL cholesterol (%), LDL cholesterol (%), Triglycerides (%), Adiponectin (?g/ mL)

#### * Measures of effect

weighted mean difference

#### Data extraction (selection and coding)

Studies meeting the following criteria were included: (1) population, overweight or obese adults, We will define overweight as a body mass index (BMI) : 25 kg/m-squared and < 30 kg/m-squared (AAFP2013; WHO 2004). Obesity will be defined by a BMI : 30 kg/m-squared (AAFP 2013; WHO 2004); (2) intervention: phentermine/topiramate monotherapy; (3) comparison: placebo; (4) outcome: weight loss, percentage of people who lost weight, waist circumference, fasting glucose, fasting insulin, HbA1c, Systolic BP, Diastolic BP, total cholesterol, high-density lipoprotein-cholesterol (HDL-C), LDL cholesterol, triglycerides, adiponectin and AEs of phentermine/topiramate; (5) design, randomized controlled trials (RCTs); (6) follow-up duration, at least 4 weeks. We excluded observational studies, pooled-analyses, trials that were not randomized and trials with no control.

#### Risk of bias (quality) assessment

Two review authors will independently assess the risk of bias in included studies by considering the following characteristics: selection bias, performance bias, detection bias, attrition bias, reporting bias, and any other biases. Disagreements will be resolved by discussion, with involvement of a third review author where necessary.

#### Strategy for data synthesis

A qualitative and quantitative analysis will be provided. The qualitative analysis will focus on study design, sample size, duration of follow-up; the characteristics of enrolled patients will be summarized. The quantitative analysis will be performed through calculation of weighted mean differences for continuous outcomes and risk ratio for the dichotomous. Heterogeneity between studies will be assessed by using $I^2$, with 50% or higher regarded as high. Publication bias will be assessed visually with funnel plots. All analyses will be two-sided and will be carried out using STATA 12.0; $p < 0.05$ will be regarded as significant.

#### Analysis of subgroups or subsets

Average estimated weight loss of Subjects in different dose subgroups; Comparison between the number of people of phentermine / topiramate group and placebo group under the same weight loss effect

#### Contact details for further information

Xiang-Guo Lei


#### Organisational affiliation of the review

Guangxi Medical University
www.gxmu.edu.cn

#### Review team members and their organisational affiliations

Mr Xiang-Guo Lei. Guangxi Medical University
Mr Chen Lai. Guangxi Medical University

#### Collaborators

Dr Xi Yang. Guangxi Medical University

#### Type and method of review

Meta-analysis, Systematic review

#### Anticipated or actual start date
15 May 2020

#### Anticipated completion date
31 December 2020

#### Funding sources/sponsors
No Funding sources/sponsors

#### Conflicts of interest

#### Language
English

#### Country
China

#### Stage of review
Review Ongoing

#### Subject index terms status
Subject indexing assigned by CRD

#### Subject index terms
MeSH headings have not been applied to this record

#### Date of registration in PROSPERO
05 July 2020

#### Date of first submission
25 May 2020

#### Stage of review at time of this submission

| Stage | Started | Completed |
|---|---|---|
| Preliminary searches | Yes | Yes |
| Piloting of the study selection process | Yes | Yes |
| Formal screening of search results against eligibility criteria | Yes | No |
| Data extraction | No | No |
| Risk of bias (quality) assessment | No | No |
| Data analysis | No | No |

*The record owner confirms that the information they have supplied for this submission is accurate and complete and they understand that deliberate provision of inaccurate information or omission of data may be construed as scientific misconduct.*

*The record owner confirms that they will update the status of the review when it is completed and will add publication details in due course.*

#### Versions

05 July 2020

PROSPERO
This information has been provided by the named contact for this review. CRD has accepted this information in good faith and registered the review in PROSPERO. The registrant confirms that the information supplied for this submission is accurate and complete. CRD bears no responsibility or liability for the content of this registration record, any associated files or external websites.
