## Supplementary material for "Efficacy and safety of phentermine/topiramate in adults with overweight or obesity: A meta-analysis and systematic review": manuscript files

**S1 Table. Search strategies.**

**Source: PubMed**

**Searched on:** April 18, 2020

**Results:** 72

| **Search** | **Query** | **Results** |
| --- | --- | --- |
| #1 | Overweight OR Obesity OR weight gain OR weight loss OR body mass index OR skinfold thickness OR waist‐hip ratio OR Abdominal Fat[Mesh] | 649104 |
| #2 | (adipos* OR obes* OR overweight* OR over weight* OR fat overload syndrom* OR overeat* OR over eat* OR overfeed* OR over feed* OR body mass ind* OR waist‐hip ratio* OR skinfold thickness* OR abdominal fat*) OR (weight AND (cyc* OR reduc* OR los* OR maint* OR decreas* OR watch* OR control* OR gain* OR chang*)) | 1233264 |
| #3 | #1 OR #2 | 1270945 |
| #4 | Phentermine AND Topiramate[Mesh] | 87 |
| #5 | (phentermine AND topiramate) OR (PHEN AND TRP) OR (PHEN AND TPM) OR VI-0521 OR Qsymia OR Qnexa | 279 |
| #6 | #4 OR #5 | 279 |
| #7 | (randomized controlled trial[pt] OR controlled clinical trial[pt] OR randomized[tiab] OR placebo[tiab] OR clinical trials as topic[mesh:noexp] OR randomly[tiab] OR trial[ti]) NOT (animals [mh] NOT (humans [mh] AND animals[mh])) | 1186247 |
| #8 | #3 AND #6 AND #7 | 72 |

**Source: Embase**

**Searched on:** April 18, 2020

**Results:** 366

| **Search** | **Query** | **Results** |
| --- | --- | --- |
| #1 | 'obesity'/exp OR 'weight change'/exp OR 'weight control'/exp OR 'weight gain'/exp OR 'weight reduction'/exp OR 'body mass'/exp OR 'waist circumference'/exp OR 'waist hip ratio'/exp OR 'skinfold thickness'/exp | 995,903 |
| #2 | weight:ab,ti AND (cyc$:ab,ti OR reduc$:ab,ti OR los$:ab,ti OR maint$:ab,ti OR decreas$:ab,ti OR watch$:ab,ti OR control:ab,ti OR chang$:ab,ti OR gain:ab,ti) OR obes$:ab,ti OR overweight:ab,ti OR 'over weight':ab,ti OR overeat:ab,ti OR 'over eat':ab,ti OR overfeed:ab,ti OR 'over feed':ab,ti OR 'fat overload syndrom$':ab,ti OR 'body mass ind$':ab,ti OR 'waist hip ratio':ab,ti OR 'waist circumferenc$':ab,ti OR adipos$:ab,ti OR 'abdominal fat':ab,ti OR 'skinfold thickness':ab,ti | 829,509 |
| #3 | #1 OR #2 | 1,392,131 |
| #4 | 'phentermine'/exp AND 'topiramate'/exp | 492 |
| #5 | phentermine:ab,ti AND topiramate:ab,ti OR (phen:ab,ti AND trp:ab,ti) OR (phen:ab,ti AND tpm:ab,ti) OR 'vi 0521':ab,ti OR qsymia:ab,ti OR qnexa:ab,ti | 425 |
| #6 | #4 OR #5 | 758 |
| #7 | 'crossover procedure':de OR 'double-blind procedure':de OR 'randomized controlled trial':de OR 'single-blind procedure':de OR random*:de,ab,ti OR factorial*:de,ab,ti OR crossover*:de,ab,ti OR ((cross NEXT/1 over*):de,ab,ti) OR placebo*:de,ab,ti OR ((doubl* NEAR/1 blind*):de,ab,ti) OR ((singl* NEAR/1 blind*):de,ab,ti) OR assign*:de,ab,ti OR allocat*:de,ab,ti OR volunteer*:de,ab,ti | 2,541,022 |
| #8 | #3 AND #6 AND #7 | 366 |

**Source: CENTRAL**

**Searched on:** April 18, 2020

**Results:** 38

| **Search** | **Query** | **Results** |
| --- | --- | --- |
| #1 | MeSH descriptor Obesity explode all trees | 13219 |
| #2 | MeSH descriptor Weight gain explode all trees | 2483 |
| #3 | MeSH descriptor Weight loss explode all trees | 6115 |
| #4 | MeSH descriptor Body mass index explode all trees | 9829 |
| #5 | MeSH descriptor Skinfold thickness explode all trees | 317 |
| #6 | MeSH descriptor Waist‐hip ratio explode all trees | 251 |
| #7 | MeSH descriptor Abdominal fat explode all trees | 478 |
| #8  #9 | MeSH descriptor Overweight explode all trees  overweight* in All Text or (over in All Text and weight* in All Text) or (fat in All Text and overloadin All Text and syndrom* in All Text) or (overeat* in All Text or (overin All Text and eat* in All Text) ) or overfeed* in All Text or (overin All Text and feed* in All Text) or adipos* in All Text or obes*in All Text or (weight in All Text near/3 cyc*in All Text) or (weight in All Text near/3 reduc*in All Text) or (weight in All Text near/3 los*in All Text) or (weight in All Text near/3 maint*in All Text) or (weight in All Text near/3 decreas*in All Text) or (weight in All Text near/3 watch*in All Text) or (weight in All Text near/3 control*in All Text) or (weight in All Text near/3 gain*in All Text) or (weight in All Text near/3 chang*in All Text) or (body in All Text and massin All Text and ind* in All Text) or (waist‐hipin All Text and ratio* in All Text) or (skinfold in All Text and thickness*in All Text) or (abdominal in All Text and fat*in All Text) | 15691  4744 |
| #10 | #1 or #2 or #3 or #4 or #5 or #6 or #7 or #8 or #9 | 29810 |
| #11 | MeSH descriptor Phentermine explode all trees | 108 |
| #12 | MeSH descriptor Topiramate explode all trees | 526 |
| #13 | (phentermine in All Text and topiramate in All Text) or (PHEN in All Text and TRP in All Text) or (PHEN in All Text and TPM in All Text) or VI-0521 in All Text or Qsymia in All Text or Qnexa in All Text | 236 |
| #14 | #11 and #12 or #13 | 256 |
| #15 | #10 and #14 | 38 |

**Source: Clinicaltrials.gov**

**Searched on:** April 18, 2020

**Results:** 33

| **Search** | **Query** | **Results** |
| --- | --- | --- |
| #1 | (phentermine AND topiramate) OR (PHEN AND TRP) OR (PHEN AND TPM) OR VI-0521 OR Qsymia OR Qnexa | 33 |
