## Supplementary material for "Efficacy and safety of phentermine/topiramate in adults with overweight or obesity: A meta-analysis and systematic review": manuscript files

Table S2 Characteristics of randomized controlled studies included in meta-analysis

| **Author** | **Year of publication** | **NCT number** | **Region** | **Study duration (weeks)** | **Dosage of drug (mg)** | **Co-intervention** | **Number of patients** | **Sex (n = Female)** | **Age (years)** | **Baseline BMI (kg/m**^2^) | **Baseline weight (kg)** |
| --- | --- | --- | --- | --- | --- | --- | --- | --- | --- | --- | --- |
| Gadde | 2011 | NCT00553787 | USA | 56 |  | standardized lifestyle counseling | Placebo (n = 994) | Placebo (n = 695) | Placebo 51.2 (10.25) | Placebo  36.7 (4.6) | Placebo  103.3 (18.1) |
|  |  |  |  |  | P/T 7.5/46 |  | P/T 7.5/46 (n = 498) | P/T 7.5/46 (n = 349) | P/T 7.5/46 51.1 (10.43) | P/T 7.5/46 36.2 (4.4) | P/T 7.5/46  102.6 (18.2) |
|  |  |  |  |  | P/T 15/92 |  | P/T 15/92 (n = 995) | P/T 15/92 (n = 693) | P/T 15/92 51.0 (10.65) | P/T 15/92  36.6 (4.5) | P/T 15/92  103.0 (17.6) |
| Winslow | 2012 | NCT00745251 | USA | 28 |  | standardized lifestyle counseling | Placebo (n = 23) | Placebo (n = 8) | Placebo 51.4 (5.74) | Placebo  35.3 (3.14) | Placebo  106.9 (16.65) |
|  |  |  |  |  | P/T 15/92 |  | P/T 15/92 (n = 22) | P/T 15/92 (n = 13) | P/T 15/92 53.4 (6.95) | P/T 15/92  36.0 (3.07) | P/T 15/92  103.7 (14.56) |
| Garvey | 2012 | NCT00796367 | USA | 108 |  | standardized lifestyle counseling | Placebo (n = 994) | Placebo (n = 695) | Placebo 51.2 (10.3) | Placebo  36.7 (4.6) | Placebo  103.3 (18.1) |
|  |  |  |  |  | P/T 7.5/46 |  | P/T 7.5/46 (n = 498) | P/T 7.5/46 (n = 349) | P/T 7.5/46 51.1 (10.4) | P/T 7.5/46 36.2 (4.4) | P/T 7.5/46  102.6 (18.2) |
|  |  |  |  |  | P/T 15/92 |  | P/T 15/92 (n = 995) | P/T 15/92 (n = 693) | P/T 15/92 51.0 (10.7) | P/T 15/92  36.6 (4.5) | P/T 15/92  103.0 (17.6) |
| Allison | 2011 | N.R. | USA | 56 |  | standardized lifestyle counseling | Placebo (n = 514) | Placebo (n = 425) | Placebo 43.0 (11.76) | Placebo  (n = 513) 42.0 (6.15) | Placebo  (n = 513) 115.8 (21.46) |
|  |  |  |  |  | P/T 3.75/23 |  | P/T 3.75/23 (n = 241) | P/T 3.75/23 (n = 201) | P/T 3.75/23 43.0 (10.96) | P/T 3.75/23  (n = 240) 42.6 (6.50) | P/T 3.75/23  (n = 240) 118.5 (21.85) |
|  |  |  |  |  | P/T 15/92 |  | P/T 15/92 (n = 512) | P/T 15/92 (n = 424) | P/T 15/92 41.9 (12.21) | P/T 15/92  (n = 511) 41.9 (6.04) | P/T 15/92  (n = 511) 115.2 (20.66) |
| Aronne | 2013 | NCT00563368 | USA | 28 |  | standardized lifestyle counseling | Placebo (n = 109) | Placebo (n = 86) | Placebo 45.0 (11.43) | Placebo  36.2 (3.94) | Placebo  100.0 (12.96) |
|  |  |  |  |  | P/T 7.5/46 |  | P/T 7.5/46 (n = 107) | P/T 7.5/46 (n = 85) | P/T 7.5/46 44.6 (11.07) | P/T 7.5/46 36.6 (3.94) | P/T 7.5/46  102.2 (16.48) |
|  |  |  |  |  | P/T 15/92 |  | P/T 15/92 (n = 108) | P/T 15/92 (n = 85) | P/T 15/92 44.6 (12.84) | P/T 15/92  35.9 (3.91) | P/T 15/92  99.3 (15.59) |
| Garvey | 2014 | NCT00600067 | USA | 56 |  | standardized lifestyle counseling | Placebo (n = 55) | Placebo (n = 32) | Placebo 49.5 (8.6) | Placebo  35.2 (5.0) | Placebo  98.1 (17.0) |
|  |  |  |  |  | P/T 15/92 |  | P/T 15/92 (n = 75) | P/T 15/92 (n = 58) | P/T 15/92 49.7 (7.5) | P/T 15/92  35.5 (4.7) | P/T 15/92  94.9 (17.9) |

Abbreviations: BMI, body mass index; P/T, phentermine/topiramate; N.R., not reported.

Note: All doses were taken once a day. All values are presented as mean (standard deviation).
